# VICTORY Protocol - VIrtual knowledge exchange in primary Care Through effective digital Online couRses for all Young people without borders and barriers

**DOI:** 10.64898/2026.07.11.26357803

**Authors:** Oisín Brady Bates, Balwani Mbakaya, Walter Cullen, Joe Gallagher

## Abstract

**Aim:** This study aims to design, implement, and evaluate a virtual exchange programme in global health and primary care, with a focus on building capacity and fostering collaboration between European and Sub-Saharan African institutions.

**Methods:** A mixed-methods approach will be adopted across three phases. First, a scoping review will synthesise evidence on the implementation, outcomes, barriers, and facilitators of virtual exchanges in global health. Second, a virtual exchange curriculum will be co-developed through surveys and focus groups with medical students and educators. Third, the programme will be evaluated using pre- and post-course surveys, and semi-structured interviews to measure changes in knowledge, global health competencies, and intercultural awareness. Quantitative data will be analysed using descriptive and regression analyses, while qualitative data will undergo reflexive thematic analysis.

**Conclusion:** This study will generate evidence on the design and impact of virtual exchanges for global health education, contributing to the development of sustainable and equitable curricula in PHC. By disseminating findings across academic, policy, and professional networks, the VICTORY project seeks to advance global collaboration, support workforce development, and promote health equity.

## Background

### 1. The importance of global primary care

#### Globalisation of health

Global health is an area of study, research, and practice that prioritizes improving health and achieving health equity for all people worldwide. The globalisation of medicine and global health care issues are of growing and significant importance. Rather than functioning in discrete national silos, humans across the world share complex interconnected social and infrastructural systems. Health system paradigms that exist in low or middle income countries (LMICs) have significant knock-on effects in HIC countries and vice versa. These health outcomes relate to infrastructure, investment and healthcare governance.

Policy decisions in HIC countries can have profound effects on health outcomes across the world. This is highlighted in a recent report from the lancet in regards to the potential of large-scale AID programme defunding (1). Policy decisions such as cutting USAID funding has been projected to result in an increased mortality rate of between 9-19 million people by the year 2030, including 4.5 million children. Notwithstanding the tragedy of preventable human death on this scale, in LMIC countries there is a clear morale imperative to endeavour to equip our health practitioners and local health systems to deal with the huge health needs of the populations effected by these cuts. In addition, in HIC countries, health care practitioners need to be equipped to deal with the ramifications of this as well.

Due to the fact that we are part of an interconnected species that share global economic, industrial and health systems, higher levels of disease and mortality in the LMIC world will also impact the HIC world. The significance of global health as an international priority has been acknowledged for some time. In 2006 Kickbusch made the case for a “European strategy on “global health”. In this article Kickbusch defines ‘global health’ to refer to ‘those health issues which transcend national boundaries and governments and call for actions on the global forces and global flows that determine the health of people (2). It requires new forms of governance at national and international levels which seek to include a wide range of actors’. He called for coordinated policy and governance strategies in order to mitigate risks relating to, among other things, poor health outcomes, pandemics, poverty and increasing migration. Twenty years later these priority areas have not lost their importance.

The COVID-19 pandemic provided us with evidence of the interconnected nature of global health issues and the shared international ramifications of common global health issues. The COVID-19 pandemic showed us that a health issue in one country can quickly become a global existential health threat (3). The threat of infectious or zoonotic disease spread across international borders, is in of itself a key global health concern. However, issues such as a global pandemic do not occur in isolation and are instead influenced by a complex web of interconnected economic, political, infrastructural and global health systems (4, 5). A worrying infectious disease is a concern that requires international cooperation and robust public primary and tertiary care health structures that are capable of preventing, monitoring and mitigating transmission. A disease like COVID-19 is influenced along the entire transmission and infection pathway by global health factors. The development of a virulent pathogen like COVID-19 is strongly impacted by global health factors like increasing urbanisation, biodiversity loss and population density (6, 7). Consequently, the spread of this pathogen was influenced by factors such as access to clean water and migrant and population health factors. For example, when a fundamental component of social infrastructure like access to clean water is disrupted due to an extreme weather event such as flooding or drought, proper hygiene becomes a major concern(8). In addition, without the appropriate public health messaging and communication, the correct public measures cannot be adopted (9, 10). This example illustrates some of the complex and interwoven direct and indirect effects relating to global health issues and their drivers, in the context of the COVID-19 pandemic. However, these lessons can be applied to all global health issues beyond the risk of infectious disease, including non-communicable disease (11), and issues relating to climate change and global warming (12). As such, there is a necessity to acknowledge global health as a priority for national and international policy makers, in addition in the education of healthcare providers.

As mentioned above, climate change and environmental instability are other examples of global health issues, with clear international consequences, that requires a coordinated international approach and awareness (13). The long-term public health benefits of carbon reduction and climate change mitigation are significant. In 2000, climate change was estimated to be responsible for 5.5 million Disability Adjusted Life Years (14) Nearly 20 years later the Inter-governmental panel on climate change (IPCC) report published in 2019 found that in the last decade the rate of sea level rise has doubled (15). Across the world, average temperatures have increased by 1 degree above pre-industrial baseline (16), and this has already resulted in a myriad of interweaving direct and indirect health effects: severe storms and floods (17), heatwaves and droughts (18),air pollution (19),new and emerging infectious diseases (20, 21) biodiversity loss (22), poor mental health (23), reduced food yields (24), freshwater depletion (25) and increasing global conflict (26) constituting but a few examples.

Issues like climate change and global pandemics as I have outlined above not only constitute severe threats to both our individual health and survival as a species. They also drive other wide ranging political issues and health-related issues like immigration (27). Population displacement, socioeconomic pressures, increasing global conflict are all inextricably woven with health systems and health outcomes. For example, climate disruptions like extreme weather events can to population displacement and migration. A mobile population due to these factors also constitutes a risk in the transmission infectious pathogens (28). In addition, residents of more resource-poor countries with less robust national infrastructure systems will be disproportionately affected in terms of the negative health effects related to a global pandemic, further driving migration. Migrant populations with higher rates of illness and disease put economic pressure on governments and policy makers. The resources needed to address these pressures diverts money from education and welfare investment, further exacerbating these issues (29).

In the face of these complex and interweaving health outcomes related to global health issuess, it is essential that we equip doctors with the skills necessary to understand and manage these problems. Rather than educating physicians with an outlook that is insular, we should strive to produce doctors who are citizens of the world; practitioners with an involved understanding of global issues and how their practice fits into global health systems.

Strategic investments in a country’s health system and in educating the workforce for sustainability brings health benefits to the people, and also contributes to good employment, economic growth and a reduction in inequalities. The United Nation’s 2030 Agenda for Sustainable Development is designed to ensure that development is not only sustainable, but also equitable, enabling access to its dividends, including health, education and employment, for everyone (30). In order to achieve this progress towards equitable and accessible health coverage countries will need a health workforce that is aligned with population and community health needs and which is capable of adjusting to the growing demand for health care driven by rapid demographic, epidemiological, economic, social and political changes (31). In particular, primary health care (PHC) can serve as a focal point for this approach. However, this requires improvements in capacity, competency development and stronger advocacy.

#### The primary care imperative

Developments in primary health care (PHC) capacity have been acknowledged as a key goal in meeting emerging global healthcare needs. As a speciality, general practice has great potential to pioneer advancements in global health policy and to advocate for and forge connections within the realm of global health (32). In her keynote address to the 2013 WONCA World Congress, former Director-General Dr Chan stated, “Family doctors have always been the bedrock of comprehensive, compassionate, and people-centred care… A health system where primary care is the backbone and family doctors are the bedrock delivers the best health outcomes, at the lowest cost, and with the greatest user satisfaction” (33).

The Astana Declaration (2018) calls for the mobilization of all stakeholders including health professionals, academia, local and international partners, agencies and funds, to focus their efforts around the main elements of PHC as defined by the World Health Organization (WHO): meeting people’s needs through comprehensive and integrated health services (including promotive, protective, preventive, curative, rehabilitative and palliative) throughout the entire life course, prioritizing primary care and essential public health functions (32). The Declaration also stresses the importance of PHC in achieving health related Sustainable Development Goals (SDGs) (34). The core medical discipline in PHC is called general practice or family medicine (FM). In high income countries (HIC) doctors train for 2-4 years to become specialist GPs or family physicians (FPs) through academic education departments and in-service training. In Europe, strong PHC led by FPs is associated with better population health, lower rates of unnecessary hospitalizations and relatively lower socioeconomic inequality (35). However, Africa has 25% of the global disease burden but only 3% of the world’s health workforce and less than 1% of the world’s health expenditure (36). In most African countries, public sector primary care and district level services involve multidisciplinary teams of nurse practitioners, midwives, clinical officers, pharmacists, community health workers and others. Medical officers in district hospitals provide a range of medical, paediatric, surgical and obstetric procedures, usually after a post-qualification 1-2 year internship period based in tertiary teaching hospitals. They also supervise nurses or clinical officers in clinics. General practitioners in most African contexts are medical officers without specialist training, but who provide private primary care services, but these are a minority. Specialists from other vertical disciplines in the public sector also provide primary care consultations in private clinics, which they have not been trained for. As a result, health systems are often fragmented, poorly coordinated and inequitable in access. There may be challenges with inadequate financing, infrastructure and workforce, poor governance, a lack of standardised and comprehensive care, with weak supply logistics for medication, equipment and others (37).

Given the close interface that general practice has with community and the local context in which it is practiced, it is a speciality that is ideally placed to act as a focal point for a collaborative international exchange as proposed by VICTORY. VICTORY would seek to help address the gap in PHC education and skills capacity through a collaborative educational exchange between institutions in Europe and Sub-Saharan Africa (SSA)

Globally, there is compelling evidence showcasing PHC’s cost-effectiveness; comprehensive PHC and FM care results in favourable health outcomes at a low cost while maintaining high user satisfaction. However, in LMICs such as in SSA, there is a lack of standardized curricula and clinical training for FM in addition to an inconsistent delivery in terms of scope and practice(38). Consequently, PHC often remains inadequately integrated into health systems, limiting employment and career development opportunities for FPs.

In an editorial in 2018, WHO Director-General Tedros Adhanom Ghebreyesus and colleagues explained how the original vision of Alma-Ata has gone “largely unfulfilled” (39). They highlighted that investing in PHC will support tangible improvements in health and well-being and drive progress towards achieving the health targets of the Sustainable Development Goals. Education and training to support this development are vital. Recent global catastrophes such as the COVID-19 pandemic and population displacement secondary to global conflict have highlighted even more the need for a strong multidisciplinary PHC workforce articulated in the Declaration of Alma-Ata and, more recently in Astana (32).

Through strong and dynamic university collaboration, the VICTORY project will work to develop a modernised and mutually beneficial collaborative PHC curriculum between Europe and Africa, capable of providing the skills, learning and cultural awareness to students and health professionals engaged in PHC practice.

#### Health equity and addressing disparities through knowledge sharing

For PHC to achieve its critical role in meeting the growing healthcare needs of underserved populations in LMICs, it must adopt novel and versatile approaches to healthcare education. Even for those PHC professionals currently trained in SSA, many will leave their respective countries in search of greater professional opportunity, resulting in a “brain-drain” within the primary care space (40). Indeed, the demand in high- and middle-income countries is expected to continue to drive health worker migration in the coming years. International mobility of health workers may bring benefits to source and destination nations and health workers themselves, but the adverse effects of migration must be mitigated. Virtual learning solutions represent one opportunity to achieve this (41). By enhancing education environments in Africa for health workers, it is possible to ensure that individuals who wish to remain in their communities but seek educational opportunities can do so without the need to migrate. In particular, for those living in remote areas with poor access to educational resources or academic institutions, virtual learning solutions represent a possible solution (42). For those who reside in other countries, the diaspora will be able to connect with colleagues worldwide through e-learning, facilitating the sharing of learning experiences and increasing the likelihood of returning to their home country, while still accessing educational opportunities. Learners that participate in PHC knowledge exchange will also gain cultural insights and increased motivation and professional opportunities from the partnerships and networks these exchanges facilitate (43, 44).

The benefits of collaborative partnerships and collaborative research projects between institutions have been identified within the literature (45, 46). Yarmoshuk et al. 2019 (47) conducted interviews with participants at 26 international universities that had partnered with four East African universities in medicine, nursing and/or public health. Advancements in research and education were found to be of the greatest value in developing these partnerships. Of note, emphasis on social responsibility and managerial support were found to be key factors in sustaining successful partnerships. When reflecting on the value of a decades long partnership between Indiana University in North-American and Moi University in East Africa, Tierney et al. 2013 (48) remarked on developments that had been made with respect to research opportunities at both institutions and improving population health and healthcare guidelines.

This notion of the mutual strengthening of research skills and the academic growth of the institutions involved in collaborative partnerships has been referenced in other similar case studies within the literature (49–51). Informed by this, a strong emphasis on collaborative research and educational growth through communication and exchange lies at the forefront of the VICTORY programme.

#### Equitable partnerships

It is important to highlight - when initiating and developing collaborative partnerships between institutions based in countries in which there are healthcare and educational infrastructure disparities - it must be ensured that partnerships are truly cooperative and equally beneficial to both parties involved. If research priorities and the partnership agenda is dictated predominantly by one partner versus the other, there is a danger that the dynamic can become exploitative or semicolonial in nature. Frameworks for mutually successful, ethical and sustainable partnerships have been offered at various points in the literature (51–53). Larkan et al. 2016 (54) designed one such framework that recommended seven equally important core concepts essential to the development of a successful global health partnership; focus, values, equity, benefit, leadership, communication and resolution.

An adherence to a framework and values that ensure equitable, mutually beneficial and sustainable partnership would be a core principle in the formation of a collaborative relationship between

### 2. Virtual exchange

#### Virtual exchange definition

Knowledge exchange is a process in which the implicit knowledge is expressed and shared in a manner that is aimed to enhance the knowledge of exchange participants (55). Virtual exchanges are knowledge exchanges that are conducted through a combination of physical and virtual platforms (56). They consist of “sustained, technology-enabled, people-to-people education programmes” between “individuals or groups who are geographically separated and/or from different cultural backgrounds” (57). Global virtual exchange is further defined as “collaborative work among individuals, teams, and organizations that spread across countries and that is enabled by technology-mediated communication” (58)

Global lockdown in the time of the COVID-19 pandemic has necessitated a far greater reliance on e-collaborative and remote learning and communication technologies.

The development and acceptance of remote collaborative and communication technologies catalysed by the COVID-19 pandemic provides an opportunity. Rather than relying on traditional methods of education and information sharing, it has been necessary to innovate and adapt. Within the realm of global health training and research, this has led to unique and valuable educational experiences (59). Didactic presentations have been replaced by often more interactive virtual learning and communication experiences (60, 61).

Digital technologies offer opportunities to improve access to health services, enhance the responsiveness of health systems to the needs of individuals and communities, and improve the delivery of various health services. Specifically, the use of e-learning provides an opportunity for individuals in remote or rural areas or who are already employed full-time to access education, including e-mentorship and virtual research networks.

#### Benefits and negatives of virtual exchange

In recent years, an increasing number of studies have looked at the benefits related to VE programmes. In the realm of healthcare, a scoping review by Bridgwood et al. 2023 (62) categorised virtual exchange outcomes as they related to the quality of experiential learning, degree of knowledge improvement, levels of social interaction, and participant satisfaction. The opportunity for participant networking afforded by VE programmes was a strong theme highlighted in their review findings, with participants valuing the opportunity to collaborate, connect and compare healthcare experiences, which contributed to greater insights when reflecting on their own practice.

Outside the healthcare field, similar VE programme benefits have been identified. Large studies such as EVALUATE (2017–2019) and EVOLVE (2018–2021) have examined how VE contributes to the development of both teacher and student competences (63–65). Findings from these studies indicate that VE fosters students’ intercultural, digital, pedagogical, and linguistic skills (65), while also enhancing teachers’ professional competences, their ability to adopt student-centred approaches, and their proficiency in facilitating VE.

Negative outcomes and barriers to learning have also been reported in regards to telecollaboration programmes, such as virtual exchange initiatives. O’Dowd and Ritter (2006) proposed that communication breakdowns in virtual exchanges can occur across four levels: the individual, the classroom, the socio-institutional context, and the interaction itself (66). They emphasised, however, that failures usually stem from a combination of interconnected factors rather than a single cause. At the individual level, elements such as learners’ intercultural competence, their background knowledge, motivation, and expectations all play a role.

One of the main challenges at the interactional level is encouraging student engagement in exchanges to move beyond the superficial. Research highlights that learners often need support to move past assumptions of cultural similarity and instead adopt a deeper level of cultural awareness and sensitivity (67–70).

On review of the literature, commonly cited issues cited by VE programmes amongst healthcare students included concerns regarding poor VE design; specifically, delivering relevant content for time-poor students, and ensuring that this content is applicable across different student contexts (62).

#### Theoretical foundations and pedagogies

The structure of virtual exchanges and their innate reliance on robust partnerships and collaborative focus necessitates strong theoretical foundations and well considered pedagogical approaches. The flexibility afforded by hybrid learning models that incorporate e-learning approaches also provides the opportunity to utilise novel pedagogies that deviate from more traditional methods. These should be chosen with a view to optimising positive outcomes and minimising negative outcomes as discussed above.

One such educational theory is that of connectivism. Connectivism (71) is a learning theory grounded in networked digital environments. It argues that knowledge resides across networks and that inherent to successful learning is the ability to navigate, connect, and grow those networks. “In connectivism, the starting point for learning occurs when knowledge is actuated through the process of a learner connecting to and feeding information into a learning community”. Kop and Hill (2008) went further, stating, “connectivism stresses that two important skills that contribute to learning are the ability to seek out current information, and the ability to filter secondary and extraneous information” (72). The connectivism framework has been taken up in the design of virtual exchanges and collaborative online international learning (COIL) programmes (73), where distributed learners co-construct knowledge across institutional and cultural boundaries. Mackness, Mak, and Williams (2010) explained that when connectivism is applied to a MOOC, the course takes on the features of a network. Learners can act independently, many different viewpoints are included, knowledge is openly shared, and new ideas arise through these connections (74). Together, these qualities facilitate the MOOCs operation as a complex, interconnected system. De Waard et al. 2011 (75) specifically emphasise the benefits of a connectivist approach in MOOCs in relation to their importance in enabling dialogue and knowledge exchange within virtual spaces, harnessing the ability of these spaces to identify new insights “patterns of meaning” which “can be formed across regions and institutions if a network of connected people comes together”.

Connectivism also relates to the rhizomatic learning theory. The rhizomatic concept is rooted in Deleuze’s philosophy, which challenges fixed institutional and pedagological structures by proposing more fluid and open-ended alternatives (76). The image of the rhizome, drawn from botany, symbolises this logic of continual branching and interconnected structures, that lead to each other in an organic fashion. The rhizomatic structure seeks to create a space where ideas and practices coexist without exclusion. This speaks to the perspective that point A and point B can both be included, rather than the existence of point B negating point A. In the virtual education space, this approach is exemplified by Cormier’s MOOC which was designed by community members and sought to address education in a way that acknowledges the uncertainty and abundance of choice brought by the internet (77). Rhizomatic theory has been utilised in other global health adjacent e-learning approaches, ones such is a virtual learning environment or MOOC for planetary health, created by Floss et al. (78). They describe this approach, in the context of planetary health, as seeking to purposefully situate the learner among the subject matter or as they describe “the student is in nature, not external to it”. In this approach they strive to increase learner investment and self-efficacy in relation to the subject matter and offer “transformative learning that engages learners emotionally, offers hope, and moves the learner towards critical thinking”.

This emphasis on learner investment described in rhizomatic approaches is paralleled in the educational theory Constructivism. “Constructivism” was first described in the context of learning theory by Vygotsky in 1978 (79), and makes the case that learners actively build their own knowledge and understanding by integrating new information with their existing experiences and reflecting on those experiences. Rather than passively receiving knowledge, students are seen as active creators of meaning. Constructivism introduced the idea that learning is mediated by social interaction, cultural tools, and the “zone of proximal development.” The “zone of proximal development” is the range of skills a learner can perform with guidance from a more knowledgeable person but not on their own, representing their potential for development. The theory of constructivism has clear application in the context of virtual exchanges, particularly in regards to global health virtual exchanges, in which international collaborations afford access to a diverse array of cultural learning opportunities. Constructivism’s utility in regards to intercultural competency has been described by Dooly 2006 (80); showing how learners co-construct knowledge and intercultural competence through authentic, technology-mediated collaboration. Viewing the achievement of Intercultural competencies through the lens of Constructivism makes the case that these skills are best established when perspectives on other cultures are constructed and assimilated into existing perceptions and cognitive frameworks, encouraging empathy and the learner’s conscious involvement in the establishment of new perceptual paradigms in relation to different cultures (81).

Virtual exchanges might also offer advancements in Intercultural competency when viewed through the lens of the intergroup contact hypotheseis, a term first used by Allport in 1958 (82). Allport’s contact theory makes the case that intergroup prejudice can be reduced through contact between members of different social groups. This concept has been subsequently supported in multiple examples throughout the literature, including in comprehensive meta-analysis (83). With the benefit of advancing intercultural competency in mind through improved availability of access to diverse social and cultural groups, the case for virtual exchanges, and in particular international virtual exchanges, can easily be made.

In order to achieve the desired competencies and outcomes available through the utilisation of the virtual exchange approach and to help ensure learner engagement, it is important to consider the optimal pedagological methods available. With respect to learner outcomes in relation to intercultural competencies (84) Sernbo et al. 2024 also echo the benefit of a constructivist approach to pedagogy in the implemention of a virtual exchange. They describe combining a co-constructivist approach with problem based learning, with the aim of “supporting students to become active subjects in shaping their education, furthering their explorative approaches by building on their existing knowledge, rather than positioning them as passive receivers”. However, based on feedback from participating students they also highlighted the importance of a significant facilitator and teacher role in solidifying learning structures and creating a space in which effective learning can be achieved. This key role of the teachers with virtual exchange pedagogy is supported elsewhere in the literature. O’Dowd et al. 2021 (43) found that implementing pedagological mentoring as a priority helped to facilitate a deeper learner engagement and acknowledgement of cultural differences amongst exchange participants, and mitigated a tendency to reflect on cultural differences in a more superficial manner. In a prior publication O’Dowd et al. 2019 define pedagogical mentoring as “the strategies and techniques that teachers use in their classes to support students’ learning during virtual exchange projects” (65).

## Methods

This study is part of a broader initiative to develop virtual exchanges in primary healthcare between European and SSA countries.

The timeline for the project is as follows: A) participant recruitment will be completed within 6 months of study start date onset B) data collection will be completed C) results are expected within 24 months.

### Study design

This study will use a mixed methods approach, combining quantitative and qualitative data to provide a comprehensive understanding of the best approach to a global health virtual exchange programme’s design, and to measure its impact.

The research will be conducted in three phases:

- Scoping review: A scoping review of virtual exchanges in global health, identifying outcomes, best practices and facilitators.
- Curriculum development and implementation of the virtual exchange programme: A virtual exchange programme will be designed, based on a needs assessment conducted through quantitative surveys and focus groups conducted with undergraduate and postgraduate medical students and educators.
- Evaluation and data collection: A mixed methods approach will be taken to measure changes in participants’ knowledge of global health and primary care, cultural competence, and attitudes toward global collaboration.

The study will generate evidence on the design, implementation, and impact of a virtual exchange in global health. Findings will include a synthesis of best practices and facilitators from existing programmes, identification of educational needs among medical students and educators, and evaluation of the exchange’s effect on participants’ knowledge of global health and primary care, cultural competence, and attitudes toward international collaboration.

#### Study context

VICTORY is a project aiming to develop a virtual exchange project between participants in Europe (Ireland and Hungary) and in Sub-Saharan Africa (Malawi and Zimbabwe). The findings of this study will be based on the research conducted at University College Dublin, which is one of the Irish institutions involved in the larger VICTORY programme. However, the VICTORY curriculum itself will be built collaboratively, based on the experience of the European, Zimbabwean and Malawian partners, with a view to developing these areas and further building capacity and strengthening partnerships between these groups. It will promote knowledge and understanding in both Europe and Africa of the context of primary healthcare delivery in these countries and indeed between European countries (Ireland and Hungary). The virtual exchange action can assist not only in collaborating and sharing experiences and expertise but also in specific online training in curriculum and role-play simulation development.

#### Theoretical underpinnings

A scoping review was selected to examine curricular frameworks, competencies, challenges, and best practices in global health virtual exchanges, as well as their role in fostering sustainable partnerships between high- and low- to middle-income countries. The review will follow JBI methodology (85), PRISMA-ScR guidelines (86), and the six-step framework of Arksey and O’Malley refined by Levac (87), covering question formulation, study selection, data charting, synthesis, and stakeholder consultation.

For phase two, relating to the design of the virtual exchange, the approach will consist of a sequential explanatory mixed methods design (88), in which the quantitative data is collected first and the focus groups are subsequently used to help explain, elaborate on, and provide a deeper understanding of the statistical results obtained in the initial quantitative phase.

The curriculum and virtual exchange design, pedagogy, and learning process will be shaped by findings from the literature review, quantitative student surveys, and focus groups with both students and educators. Expert curriculum leaders will also collaborate in developing the final content. The efficacy of the virtual exchange itself will be evaluated through pre-and post-questionnaires and qualitative semi-structured interviews. As such, a mixed methods approach will be adopted when considering both the virtual exchange design and its evaluation.

In regards to the third phase, a concurrent triangulation mixed methods design approach will be adopted, in which both the qualitative and quantitative data will be collected and analysed in tandem. Triangulation refers to the synthesising of data gleaned from both qualitative and quantitative data with a view to identifying new insights or complementary themes (89).

This study used a pragmatic framework to justify a mixed methods approach. Pragmatism, a common basis for mixed methods research (90), stresses choosing methods that best answer the research question and recognises knowledge as context-dependent and evolving (91). It is especially suited to evaluating complex, subjective issues such as attitudes and cultural awareness, where relying on one perspective may be insufficient (92).

##### Phase 1: Scoping review

The protocol for the scoping review has been published separately (93).

The central question guiding this scoping review is: How have virtual exchanges been implemented and evaluated in global healthcare education, and what are the reported outcomes, barriers, and facilitators?

This review will analyse pedagogical frameworks, competencies, and challenges in global health virtual exchanges, focusing on their potential to strengthen sustainable partnerships. It will also identify gaps in the literature to inform curriculum design.

##### Phase 2: Implementation of the virtual exchange programme

A multidisciplinary team of experts in global health and primary care technology will design the virtual exchange curriculum in collaboration with students and trainees. The curriculum will focus on core concepts in global health and primary care, with case studies from different countries, collaborative projects, and synchronous/asynchronous discussions.

A needs assessment will be conducted through quantitative surveys and qualitative focus groups to aid in determining design and learning approaches.

The virtual exchange will be delivered over a 12-week period, with structured learning activities, virtual discussions and reflective exercises.

#### Study design and objectives

Data will be gathered via a mixed methods approach.

The first phase of the study will consist of a questionnaire, disseminated to all undergraduate and postgraduate medical students at UCD. The second phase of this study will consist of a series of semi-structured focus group interviews with students and educators at UCD.

The objectives of the mixed methods study are:

1. To explore the perceptions and experiences of students and educators participating in the virtual exchange;
2. To conduct a needs assessment in regards to current primary care and global health training;
3. To evaluate perceived best pedagogical approaches;
4. To identify barriers and facilitators to the sustainability and scalability of virtual exchange programs in medical education.

The objectives of the survey are:

5. To explore the perceptions and experiences of students and educators participating in the virtual exchange;
6. To conduct a needs assessment in regards to current primary care and global health training;
7. To evaluate perceived best pedagogical approaches;
8. To identify barriers and facilitators to the sustainability and scalability of virtual exchange programs in medical education.

The overarching aim of the focus groups is: To clarify the information and data necessary for developing the course materials for virtual exchanges focusing designated curriculum topics and to gather insights from third-level students and educators participating in a virtual exchange programme on what they would like to learn and how they would like to learn it.

The specific objectives of the focus groups are:

1. To ascertain curricular learning objectives and topics of interest, from a participant perspective;
2. To identify participants preferred learning methods and tools;
3. To explore participant preferences on learner assessment methodologies;
4. To evaluate strategies for optimal learner engagement with the proposed virtual exchange programme.

#### Study population

This questionnaire will be disseminated via email to all undergraduate and postgraduate medical students at UCD. An invitation will be extended to all undergraduate and postgraduate medical students currently enrolled at UCD, Dublin. This corresponds to approximately 1000 students. The anticipated uptake rate for students will be at least 25% therefore an approximated sample size will be 250 students.

There will be approximately 5 focus groups held. 20-25 students will be involved in four of these focus groups and approximately 10-15 educators will be invited to participate in the last of these 5 groups. Staff from the UCD school of medicine will be eligible to participate. Purposive sampling will be used in relation to the educator cohort; study participants will be chosen based on their specialty, level of responsibility and gender.

#### Data collection

An e-mail inviting students to participate in the study will be sent to all undergraduate year groups using e-mail class lists. The e-mail invite will contain an information leaflet which will gave details about the study overall and survey and a brief introductory note summarizing the purpose of the study and the contents of the survey. The e-mail invite also contained a link to a secure website (https://www.surveymonkey.com/) (94) for completing the anonymous survey.

Questions are grounded in frameworks such as Kirkpatrick’s Training Evaluation Model (which focuses on participant reaction, learning, behaviour, and results) (95), Moore’s Outcomes Hierarchy (which focuses on healthcare professional education outcomes) (96), and Miller’s Pyramid of Clinical Competence (which addresses stages of knowledge and performance in clinical settings) (97). Similar studies evaluating medical students’ needs and curriculum perceptions can be found in literature such as Cook et al.’s reviews on technology-enhanced learning in medical education (98).

The semi-structured focus groups topics and question schedules were guided by the findings from the questionnaire and literature review. A structured questionnaire template will be developed for each focus group topic however the questions in these templates were followed up with prompts in order to encourage free flowing conversation and organic participation. Prior to initiating the formal semi-structured interviews, a pilot interview will be performed.

#### Data analyses

##### Questionnaire

Data will be exported from the surveymonkey.com website into excel documents. Data will be fully anonymised by the surveymonkey.com online survey-tool prior to exporting the data into Excel.

The data will then then cleaned and imported into SPSS or a similar statistical analysis tool in order to analyse trends and compare the responses in relation to various characteristics and between different groups. The “Exploration” descriptive statistics tool in SPSS will be employed to demonstrate trends and tendencies within the data. Comparisons in responses between groups will be investigated using ordered logistic regression.

Help will enlisted from the statistics office, UCD in the analysis and interpretation of the data.

##### Focus groups

Braun and Clarke’s (2006) (99) framework for thematic analysis will be employed in the analysis of the semi-structured focus groups. Reflexive thematic analysis was deemed the most suitable analytical method in this study as it encourages a flexible and organic coding process. The reflexive thematic process endorsed by Braun and Clarke facilitates the conceptualization of themes as reactive and determined by the data. It allows forensic analysis of the data and the identification of shared themes and ideas which are underpinned by an overlying concept. In this way it supports the reliability of qualitative coding through a robust, iterative reflection on the data and stimulates original and multifaceted findings that are independent from the study aims and objectives which have been determined from the outset.

###### Phase 3 - Virtual exchange evaluation

The next phase of the study involved evaluation of the virtual exchange programme.

#### Study design and objectives

##### Quantitative Component

- Pre- and post-surveys will be used to assess changes in participants’ knowledge of global health and primary care, cultural competence, and attitudes toward global collaboration.

- Learning analytics (e.g., engagement with the platform, completion of tasks) will be collected to assess participation and engagement levels.

##### Qualitative Component

- Semi-structured interviews will be conducted with participants and educators to explore their experiences, challenges, and perceptions of the virtual exchange.

- Content analysis will be used to examine themes related to cultural learning, collaboration, and global health perspectives.

The objectives of this phase of the third phase of the study are:

1. To measure advancements in global health and primary care competencies amongst course users
2. To assess participant satisfaction with the virtual exchange course and overall experience
3. To evaluate changes in learner intercultural competencies and awareness
4. To make recommendations for the future and ongoing use of virtual exchange programmes in global and primary care

#### Study population and sampling

For the quantitative component, the study will recruit all users of the virtual exchange curriculum materials, at UCD. Based on similar virtual exchange programme experiences (100), the expected participation number is between 30-80.

In regards to the semi-structured interviews, purposive sampling will be employed in order to ensure a diverse array of respondents with varied demographic characteristics. In relation to educators, an effort will be made to sample participants across a broad range of specialties and from varying levels of responsibility and seniority within the delivery of curriculum. Every effort will be made to ensure a balance with respect to gender.

##### Data collection

Data will be collected using an inbuilt questionnaire/survey money tool integrated into the online curriculum materials. Post course questionnaires will be emailed to all course participants. The completion of both pre and post-course questionnaires will be mandatory to receive certification of course completion.

Interview and focus group participants will be invited to participate via email. Interviews and focus groups will be conducted over a video communication platform. All participants and the interviewer will be required to have their camera turned on so that they can maintain face-to-face contact throughout the interview via video link. Nobody else will be present during the interviews apart from the principal researcher and the participants.

Interviews will last from 30 minutes to 1 hour. When data saturation is achieved, two further interviews will be conducted and one further focus group will be conducted. The definition of data saturation is taken from the description given by Grady 1998 (101).

##### Data analyses

###### Quantitative Analysis

To evaluate the outcomes and experiences associated with the primary care global health virtual exchange program, we conducted a quantitative assessment using structured, self-administered questionnaires aligned with Kirkpatrick Levels 1 and 2 (102). These tools were adapted from validated instruments and recent research on equitable global health education (103, 104), with additional reference to the Intercultural Sensitivity Scale (105) and the 21^st^ century skills framework (70, 106).

###### Pre-Course Assessment

Prior to course commencement, all participants will complete:

- A Participant Characteristics Questionnaire to gather sociodemographic and contextual data. Based on similar research in this area (78), and from questions asked quantitative surveys to students in the first phase of the programme’s research when investigating student perspectives on the programme design.
- Questions based on the “21^st^ century skills”, used elsewhere to measure competency development in virtual exchange programmes.
- A Global Primary Care Competency Self-Assessment. The questions in the self-assessment were modelled off the global health competency domains identified by Walpole et al. 2016 (from CUGH domains) (103, 107) and with adaptations taken from the global health competency questions used in Alayande et al. (2023) (104).

###### Post-Course Assessment

At course completion, participants completed a suite of questionnaires to evaluate:

- Change in Global Primary Care Competencies (Kirkpatrick Level 2: Learning – post) and the 21^st^ century skills framework (95, 106).
- Course Evaluation and Satisfaction (Kirkpatrick Level 1: Reaction), including structure, engagement, and applicability of content (95).
- Perceptions of Small Group Dynamics, exploring collaborative learning, cultural exchange, and team functionality, adapted from Alayande et al. (2023) (104).
- Online Learning Experience, addressing digital platform usability, learning environment, and perceived disadvantages of virtual participation.
- Intercultural Sensitivity Scale (ISS), a 44-item validated measure of participants affective and cognitive orientation toward intercultural interaction(105).

In addition, a faculty-specific survey will be administered post-course to explore equity in course design and delivery. This instrument is informed by the Fair Trade Learning rubric and probes used to assess equity of institutional partnerships in Alayande et al. (2023) (104).

All instruments will be hosted on secure online forms via the Surveymonkey tool, and participation will be voluntary following informed consent. For analysis, data will be exported from the surveymonkey.com website into excel documents. Data will be fully anonymised by the surveymonkey.com online survey-tool prior to exporting the data into Excel.

The data will then then cleaned and imported into SPSS or a similar statistical analysis tool in order to analyse trends and compare the responses in relation to various characteristics and between different groups. The “Exploration” descriptive statistics tool in SPSS will be employed to demonstrate trends and tendencies within the data. Comparisons in responses between groups will be investigated using ordered logistic regression.

Help will be enlisted from the statistics office, UCD in the analysis and interpretation of the data.

##### Qualitative analysis

###### Post programme Focus group – Intercultural competencies developed by users during the course

As part of a mixed-methods evaluation of user experiences in the primary care global health virtual exchange program, a series of post-course focus groups will be conducted.

One of these focus groups will explore participant; intercultural experiences and development through the lens of the Council of Europe’s Competences for Democratic Culture (CDC) framework (108).

The CDC framework provides a comprehensive set of competences essential for individuals to participate effectively in culturally diverse democratic societies. Drawing on O’Dowd’s (2021) application of this framework in virtual exchange contexts, our focus group design will explore participants’ reflections across key CDC domains (43). These include openness to cultural otherness, critical self-awareness, cultural knowledge, empathy, cooperation, conflict-resolution, and digital communication competence.

Focus group discussions will be semi-structured, with prompts aligned to the CDC domains and adapted from O’Dowd’s coding framework. Participants will be invited to reflect on how the virtual exchange shaped their intercultural awareness, engagement with global health challenges, and ability to work collaboratively across diverse sociocultural contexts.

Transcripts will be coded thematically using a deductive approach based on CDC domains, alongside inductive coding to capture unanticipated themes. This approach will help identify specific intercultural competencies developed through the exchange, supporting evaluation of the program’s broader educational impact.

###### Post-programme Semi-Structured interviews

Semi-structured interviews will also be performed following the virtual exchange component with educators and users. These interviews will explore the experience of the virtual exchange and associated challenges. The semi structure interview guide will be based on findings from Protsiv et al., Alayande et al and the Kirkpatrick levels (95, 104, 109).

## Discussion

### Strengths and limitations

The mixed-methods approach adopted by this study is a key strength, ensuring a comprehensive understanding of global health virtual exchange best practices. This study will be the only study evaluating both student and educator perspectives on global health virtual exchanges, to the best of the principal researcher’s knowledge.

However, there are some limitations to the study which might constraints on the research and limit the applicability of its findings. The researchers involved in this study all also have roles as educators at UCD, therefore when sampling educators and conducting focus groups or interviews, there is a risk of bias, as the participant’s relationship with the researchers may influence their responses to the questions posed. In order to eliminate bias, participants will be explicitly advised as to the anonymisation of data. Furthermore, the interviewer will endeavour to remain as neutral as possible when both asking questions and responding to participant statements.

Furthermore, a proportion of students may be familiar with the researchers due to previous teaching interactions. This familiarity may influence student uptake in regard to the quantitative surveys. In order to mitigate this, in the medical student invite e-mail and information leaflet, participants will be advised that all their responses will be entirely anonymised, reducing the likelihood that the participant’s former knowledge of the author would provoke bias when inputting survey responses.

### Discussion

The findings of this study have important implications for dissemination and scalability. Dissemination should occur through peer-reviewed publications, academic conferences, and open-access repositories to ensure broad reach across both high- and low-income settings. Equally, engagement with professional networks such as WONCA, the European General Practice Research Network, and African academic consortia will be key to embedding outputs into curricula and policy dialogues. Beyond academic settings, targeted dissemination to ministries of health and education will be essential to align curricular innovations with workforce development strategies.

Future directions include iterative refinement of the virtual exchange model based on evaluation data, with attention to sustainability and equitable partnership structures. Expansion to additional institutions across Sub-Saharan Africa and Europe will allow comparative analyses of context-specific outcomes and facilitate long-term capacity building. Integration with digital platforms that support interactivity and longitudinal mentoring should also be prioritised to strengthen participant engagement and knowledge retention.

In conclusion, this study highlights the potential of virtual exchanges to enhance global health and primary care education, strengthen intercultural competencies, and support equitable academic partnerships. By embedding rigorous evaluation and dissemination strategies, the VICTORY project can contribute meaningfully to addressing disparities in training and to shaping a more globally connected primary care workforce.

## Ethical Considerations

Ethical approval will be sought from relevant institutional review boards (IRBs) in both participating countries. Informed consent will be obtained from all participants, and data will be anonymized to protect privacy. Participants will have the right to withdraw from the study at any time.

## Author Contributions

Oisín Brady Bates: Conceptualisation, Methodology, Writing – Original Draft, Writing – Review & Editing, Project Administration

Balwani Mbakaya: Conceptualisation, Writing – Review & Editing

Walter Cullen: Conceptualisation, Writing – Review & Editing

Joe Gallagher: Conceptualisation, Methodology, Writing – Review & Editing, Project administration

## Data Availability Statement

There is no publicly available date related to this protocol.

